# Combination therapy with tocilizumab and corticosteroids for aged patients with severe COVID-19 pneumonia: a single-center retrospective study

**DOI:** 10.1101/2020.09.26.20202283

**Authors:** Francisco López-Medrano, María Asunción Pérez-Jacoiste Asín, Mario Fernández-Ruiz, Octavio Carretero, Antonio Lalueza, Guillermo Maestro de la Calle, José Manuel Caro, Cristina de la Calle, Mercedes Catalán, Rocío García-García, Joaquín Martínez-López, Julia Origüen, Mar Ripoll, Rafael San Juan, Hernando Trujillo, Ángel Sevillano, Eduardo Gutiérrez, Borja de Miguel, Fernando Aguilar, Carlos Gómez, José Tiago Silva, Daniel García-Ruiz de Morales, Miguel Saro-Buendía, Ángel Marrero-Sánchez, Guillermo Chiara-Graciani, Héctor Bueno, Estela Paz-Artal, Carlos Lumbreras, José L. Pablos, José María Aguado, on behalf of the H12O Immunomodulation Therapy for COVID-19 Group

**Author notes:** ***Corresponding author***: Francisco López-Medrano, MD, PhD, MSc. Unit of Infectious Diseases. Hospital Universitario “12 de Octubre”. Centro de Actividades Ambulatorias, 2^a^ planta, bloque D. Avda. de Córdoba, s/n. Postal code 28041. Madrid, Spain. These authors equally contributed to the present work. Other members are listed in the Appendix.

## Abstract

**Background:** The role of combination immunomodulatory therapy with systemic corticosteroids and tocilizumab (TCZ) for aged patients with COVID-19-associated cytokine release syndrome remains unclear.

**Methods:** We conducted a retrospective single-center study including consecutive patients ≥65 years that developed severe COVID-19 between March 3 and May 1, 2020 and were treated with corticosteroids at various doses (methylprednisolone [0.5 mg/Kg/12 hours to 250 mg/24 hours]), either alone (“CS group”) or associated to intravenous tocilizumab (400-600 mg, one to three doses) (“CS-TCZ group”). Primary outcome was all-cause mortality by day +14, whereas secondary outcomes included mortality by day +28 and clinical improvement (discharge and/or a ≥2-point decrease on a six-point ordinal scale) by day +14. Propensity score (PS)-based adjustment and inverse probability of treatment weights (IPTW) were applied.

**Results:** Overall, 181 and 80 patients were included in the CS and CS-TCZ groups. All-cause 14-day mortality was lower in the CS-TCZ group, both in the PS-adjusted (hazard ratio [HR]: 0.34; 95% confidence interval [CI]: 0.17 – 0.68; *P-*value = 0.002) and IPTW-weighted models (odds ratio [OR]: 0.38; 95% CI: 0.21 – 0.68; *P*-value = 0.001). This protective effect was also observed for 28-day mortality (PS-adjusted HR: 0.38; 95% CI: 0.21 – 0.72; *P*-value = 0.003). Clinical improvement by day +14 was higher in the CS-TCZ group in the IPTW analysis only (OR: 2.26; 95% CI: 1.49 – 3.41; *P*-value <0.001). The occurrence of secondary infection was similar between both groups.

**Conclusions:** The combination of corticosteroids and TCZ was associated with better outcomes among patients ≥65 years with severe COVID-19.

## Introduction

Severe acute respiratory syndrome coronavirus 2 (SARS-CoV-2), the causative agent of Coronavirus Disease 2019 (COVID-19), is a novel beta-coronavirus first identified in December 2019 in Wuhan, China (1). Most cases of SARS-CoV-2 infection have a mild-to-moderate course, although a significant proportion of patients will ultimately develop acute respiratory distress syndrome (ARDS) (2), which carries a high mortality rate (3). Older age has been consistently identified as a risk factor for death and poor outcomes in COVID-19. For example, a report of 72,314 cases from China found case-fatality rates of 8% and 15% for patients aged 70-79 years and >80 years, respectively, in contrast with an overall rate as low as 2.3% for the global cohort (4). In the United Kingdom the risk of death for COVID-19 patients older than 80 years was 20-fold higher than those between 50 and 59 (5).

The pathogenesis of severe COVID-19 and associated ARDS involves a hyperinflammatory status resembling the cytokine storm release syndrome observed in patients with influenza or immune-mediated diseases (6). Therefore, a growing number of immunomodulatory therapies are being tested to counteract this deleterious inflammatory response (7-15). One of the most mature approaches at this point of the pandemic involves the off-label use of the humanized anti-interleukin (IL)-6 receptor monoclonal antibody tocilizumab (TCZ) (9-11), on the basis of the previous experience with patients receiving chimeric antigen receptor T-cell therapy (16). On the other hand, a recently published randomized controlled trial (RCT) has demonstrated a survival benefit from corticosteroid therapy in patients with ARDS (17). In the setting of COVID-19, a meta-analysis comprising more than 1,700 participants from 7 RCTs reported a reduction in 28-day mortality of 34% with the administration of systemic corticosteroids as compared with usual care or placebo(18).

Regardless this emerging evidence, studies comparing both TCZ- and corticosteroids-based therapeutic strategies are scarce (19). The additional benefit to be expected from the sequential use of TCZ in the context of previous use of corticosteroids has been recently raised (20). This question is particularly relevant for older patients, which have an increased risk of death due to COVID-19 but also of developing adverse events related to immunomodulatory agents, such as superinfections. The shortage of TCZ during the first weeks of the pandemic in our setting allowed us to perform a comparative study focused on the vulnerable population of individuals aged 65 years and older with the objective to analyze the value of adding TCZ to systemic corticosteroid therapy (as compared to corticosteroids alone) in patients with severe COVID-19 pneumonia.

## Materials and Methods

### Study population and design

This retrospective cohort study was conducted at the University Hospital “12 de Octubre” a 1,300-bed tertiary care center located in the urban area of Madrid (Spain). The Clinical Research Ethics Committee approved the study protocol and granted a waiver of informed consent due to its observational design. The research was performed in accordance with the Strengthening the Reporting of Observational Studies in Epidemiology (STROBE) guidelines. The study population included patients ≥65 years consecutively admitted due to severe COVID-19 pneumonia from March 3 to May 1, 2020, that received intravenous (IV) corticosteroids alone (“CS group”) or associated to IV TCZ (“CS-TCZ group”) as immunomodulatory therapy. For analysis purposes, the date of administration of the first dose of any of these agents was considered as day 0. To minimize potential survivor bias, patients that died within the first 24 hours (i.e. day +1) were excluded. Participants were followed-up to discharge, death (whichever occurred first), or June 30, 2020.

Demographics and comorbidities; symptoms at presentation; vital signs, laboratory values and radiological features at day 0; use of antiviral therapy; treatment-related adverse events; and outcomes were collected from electronic medical records using a standardized case report form. Respiratory function was assessed by the pulse oximetry oxygen saturation/fraction of inspired oxygen (SpO_2_/FiO_2_) ratio. The National Early Warning Score (NEWS) was calculated at admission. Dynamic changes over time in the clinical status were assessed according to a six-point ordinal scale: 1.- discharged to home; 2.- admitted to the hospital, not requiring supplemental oxygen; 3.- admitted to the hospital, requiring low-flow supplemental oxygen (FiO_2_<40%); 4.- admitted to the hospital, requiring high-flow supplemental oxygen (FiO_2_ ≥40%) or non-invasive mechanical ventilation; 5.- admitted to the hospital, requiring invasive mechanical ventilation (IMV), extracorporeal membrane oxygenation (ECMO), or both; and 6.- death.

### Antiviral and immunomodulatory therapies

According to clinical practice guidelines issued by the Spanish Ministry of Health during the study period (21), antiviral regimens could include co-formulated lopinavir/ritonavir (LPV/r) (200/100 mg twice daily for up to 14 days), hydroxychloroquine (HCQ) (400 mg twice for the first day, followed by 200 mg twice daily for 5-10 days), and subcutaneous (SC) interferon (IFN)-β (250 μg every 48 hours). All these drugs were used off-label and oral or written informed consent was previously obtained. In addition, some patients received IV remdesivir (200 mg during the first day, followed by 100 mg daily for 5 to 10 days) in the context of an ongoing clinical trial. The use of empirical antibiotic therapy was contemplated for patients admitted due to COVID-19 pneumonia. Most patients received thromboprophylaxis with low-molecular-weight heparin.

Due to shortages in TCZ supply, a multidisciplinary committee that included clinical specialties and the Department of Pharmacy was created to facilitate and standardize therapeutic decisions, as detailed elsewhere. The committee held daily meetings (except for the weekends) throughout the pandemic period. The first meeting took place on March 18, two days after the introduction in the local protocol of TCZ as a therapeutic option for COVID-19. The off-label use of TCZ was considered for patients potentially eligible for intensive care unit (ICU) admission, with bilateral or rapidly progressive infiltrates in chest X-ray or computerized tomography (CT) scan, and fulfilling one or more of the following criteria: respiratory rate >30 bpm and/or pulse oximetry oxygen saturation (SpO_2_) <92% on room air, C-reactive protein (CRP) level >10 mg/dL, IL-6 level >40 pg/mL, and/or D-dimers >1,500 ng/mL. Exclusion criteria included the presence of liver function abnormalities (alanine aminotransferase and/or aspartate aminotransferase levels >5 times the upper normal limit), uncontrolled bacterial or fungal infection, or acute diverticulitis or bowel perforation. An initial IV 400 mg (if body weight <75 Kg) or 600 mg (if body weight ≥75 Kg) dose was administered as one-hour infusion. Until March 26, a second 400 mg dose was routinely administered 12 hours later, whereas a third dose could be given after 24 hours from the first infusion for selected patients according to the criteria of the treating physician (22). After that date, a single TCZ dose was administered according to the updated recommendations of the Ministry of Health of Spain, on the basis of dosing regimens approved for rheumatologic diseases (23).

The local protocol took into account the use of corticosteroids already in its initial version (issued on March 16) for patients with severe COVID-19 pneumonia fulfilling the same inclusion criteria established for the initiation of TCZ, when the latter agent was not available or in the presence of any of the exclusion criteria detailed above. Different dosing regimens were considered (IV methylprednisolone 0.5-1 mg/Kg daily for ≤5 days or as pulses of 100 to 250 mg for 3 days). Beginning in April 2020, the prescription of corticosteroids was generalized for patients presenting to the Emergency Room with COVID-19 pneumonia and SpO2 <92% on room air, regardless of the subsequent administration of TCZ.

### Design of study groups

Given the relatively low baseline prescription rate and the marked surge in demand early after the beginning of the pandemic, there was a shortage of TCZ during the first weeks of March. The interdisciplinary committee decided to distribute, on a daily basis, the available doses according to three principles: a) the fulfillment of the previously detailed inclusion and exclusion criteria; b) the absence of a short-term life-threatening condition; c) and, providing that both previous criteria were met, the drug was preferably administered to those patients with a longer life expectancy. As a result, during the month of March TCZ was more commonly prescribed to younger patients, whereas corticosteroids alone were widely used for patients ≥65 years. As the incidence of SARS-CoV-2 infection declined and the supply of TCZ increased, an increasing proportion of patients ≥65 years received this immunomodulatory drug from April, 2020.

This circumstance allowed us to compare two cuasi-contemporary cohorts of aged patients treated with different immunomodulatory regimens: corticosteroids alone during the first period (March, 2020 [“CS group”]), and corticosteroids associated to TCZ during the second period (April and May, 2020 [“CS-TCZ group”]).

### Study outcomes

The *primary outcome* was all-cause mortality by day +14 after the first dose of the immunomodulatory therapy. S*econdary outcomes* included all-cause mortality by day +28 and clinical improvement by day +14 (defined as discharge to home and/or a decrease of ≥2 points from day 0 on the six-point ordinal scale).

### Statistical analysis

Quantitative data were shown as the mean and standard deviation (SD) or the median with interquartile range (IQR), whereas qualitative variables were expressed as absolute and relative frequencies. Categorical variables were compared using the χ^2^ test. Student’s t-test or Mann-Whitney U test were applied for continuous variables, as appropriate. Baseline factors predicting clinical improvement by day +14 were analyzed by means of logistic regression, with associations expressed as odds ratios (ORs) with 95% confidence intervals (CIs). Collinearity among explanatory variables was assessed with the variance inflation factor (VIF). Survival probabilities were graphically depicted according to the Kaplan-Meier method and compared with the log-rank test. A landmark analysis restricted to survivors that remained hospitalized by day +4 was performed to take into account potential survivor bias. Cox regression with backward stepwise selection was used to assess the effects of different variables on mortality rates by days +14 and +28. Associations were expressed as hazard ratios (HR) and 95% CIs. To overcome the limitation posed by the nonrandomized design of the study, we calculated the propensity score (PS) for receiving TCZ therapy in association to corticosteroids (versus corticosteroids alone) given the patient’s characteristics. This PS was constructed using a non-parsimonious logistic regression, with the inclusion in the CS or CS-TCZ groups considered as the dependent variable and all potential confounders entered as covariates. The ability of the resulting PS to predict the observed data was calculated by means the area under the receiver operating characteristic curve (auROC), and calibration was assessed with the Hosmer-Lemeshow goodness-of-fit test. The PS was applied in two ways to correct for baseline disparities between groups: as a covariable for regression adjustment, and as inverse probability of treatment weights (IPTW). Statistical analysis was performed with SPSS version 22.0 (IBM Corporation^®,^ Armonk, NY).

## Results

### Clinical characteristics of study groups

Overall, 275 patients ≥65 years admitted to our center with severe COVID-19 pneumonia during the study period received systemic corticosteroids. Eighty-one of them were also sequentially treated with TCZ after a median interval of 1 day (IQR: 0 - 5). Fourteen patients that died within the first 24 hours from the initiation of immunomodulatory therapy were excluded. Therefore, 181 and 80 patients were included within the CS and CS-TCZ groups, respectively.

The mean age of the global cohort was 77.2 ± 7.7 years, 147 patients (56.3%) were males, and 240 (92.0%) of Caucasian ethnicity. The median Charlson comorbidity index was 4.0 (IQR: 3 - 5). Regarding symptoms at presentation, cough was reported by 198 patients (75.9%), fever by 187 (71.6%), and dyspnea by 180 (69.0%). Among 238 patients with available data, median NEWS at admission was 4 (IQR: 2 - 6). Radiological lung involvement was diffuse in 179 patients (68.6%). All-cause 14- and 28-day mortality rates were 37.2% (97/261) and 41.4% (108/261), respectively, whereas clinical improvement by day +14 was observed in 38.3% (100/216) of patients.

We compared the clinical characteristics of patients receiving systemic corticosteroids alone or in association to TCZ. As expected in view of the evolving prescription practices over time in our center, the majority of those within the CS group were treated during the month of March (97.2% [176/181]), whereas most of patients in the CS-TCZ group were treated during April and May (92.5% [74/80]). Patients in the CS group were significantly older and had a higher prevalence of comorbid conditions, mainly obesity, chronic lung and heart disease and dementia. The NEWS at admission was also higher in this group (**Table 1**). Regarding clinical status at day 0, patients in the CS-TCZ group had a poorer respiratory status (as assessed by the SpO_2_/FiO_2_ ratio), lower lymphocyte counts and higher lactate dehydrogenase (LDH) levels, and presented more commonly with bilateral alveolar involvement in the chest X-ray. The use of LPV/r and IFN-β was more common in the CS group, whereas patients in the CS-TCZ group were more likely to be treated with remdesivir (**Table 2**).

**Table 1.**
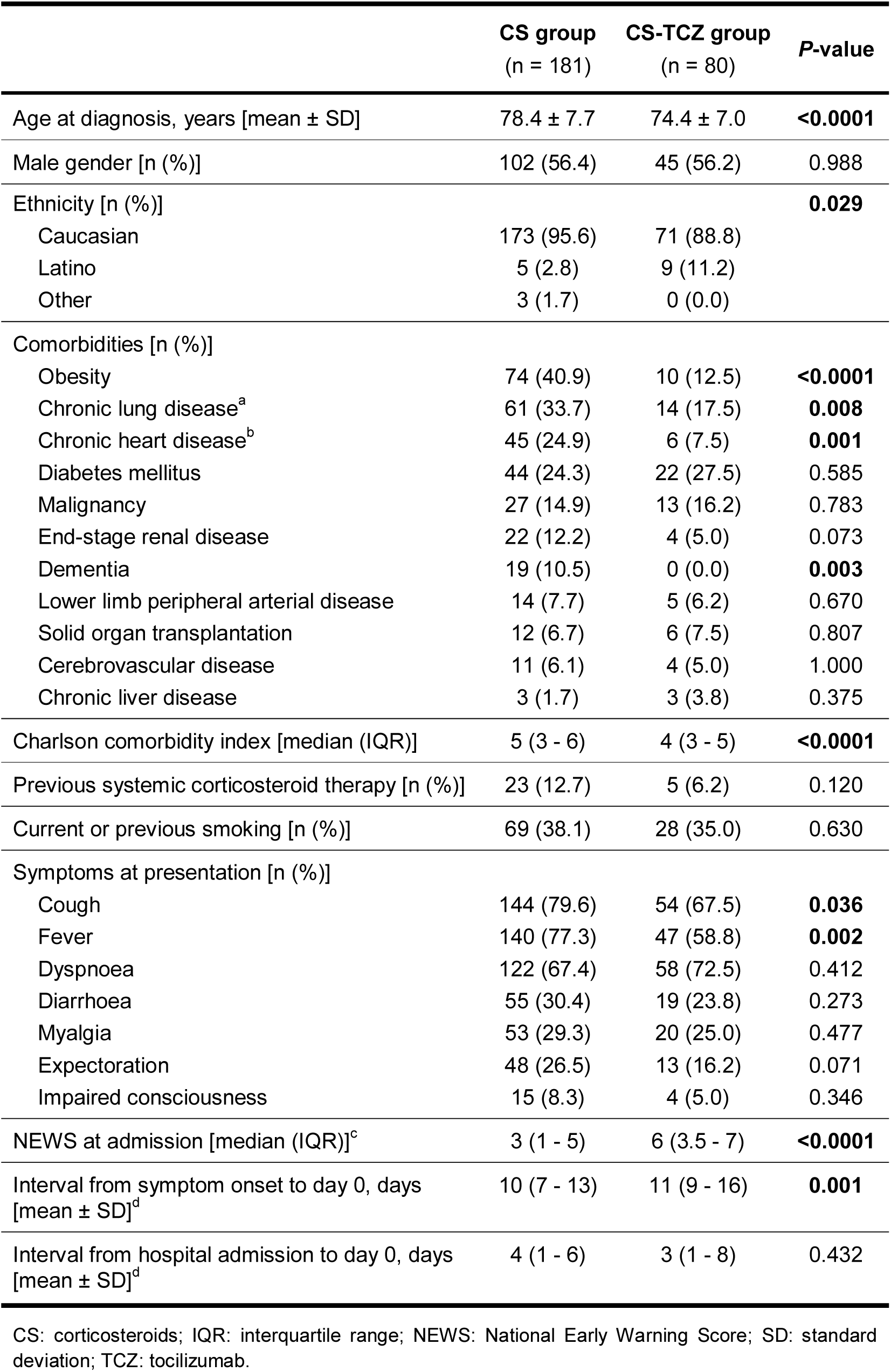
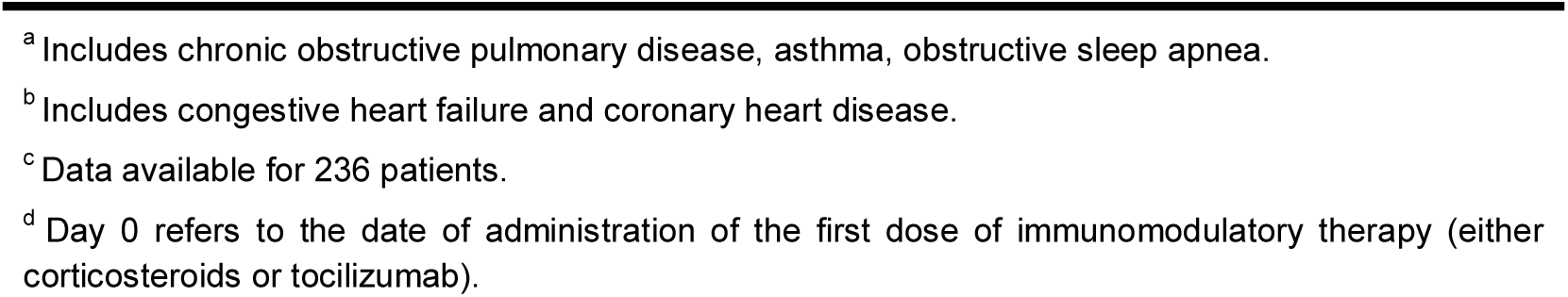
Demographics and clinical in both study groups.

|  | CS group<br>(n = 181) | CS-TCZ group<br>(n = 80) | P-value |
| --- | --- | --- | --- |
| Age at diagnosis, years [mean ± SD] | 78.4 ± 7.7 | 74.4 ± 7.0 | **<0.0001** |
| Male gender [n (%)] | 102 (56.4) | 45 (56.2) | 0.988 |
| Ethnicity [n (%)] |  |  | **0.029** |
| Caucasian | 173 (95.6) | 71 (88.8) |  |
| Latino | 5 (2.8) | 9 (11.2) |  |
| Other | 3 (1.7) | 0 (0.0) |  |
| Comorbidities [n (%)] |  |  |  |
| Obesity | 74 (40.9) | 10 (12.5) | **<0.0001** |
| Chronic lung disease<sup>a</sup> | 61 (33.7) | 14 (17.5) | **0.008** |
| Chronic heart disease<sup>b</sup> | 45 (24.9) | 6 (7.5) | **0.001** |
| Diabetes mellitus | 44 (24.3) | 22 (27.5) | 0.585 |
| Malignancy | 27 (14.9) | 13 (16.2) | 0.783 |
| End-stage renal disease | 22 (12.2) | 4 (5.0) | 0.073 |
| Dementia | 19 (10.5) | 0 (0.0) | **0.003** |
| Lower limb peripheral arterial disease | 14 (7.7) | 5 (6.2) | 0.670 |
| Solid organ transplantation | 12 (6.7) | 6 (7.5) | 0.807 |
| Cerebrovascular disease | 11 (6.1) | 4 (5.0) | 1.000 |
| Chronic liver disease | 3 (1.7) | 3 (3.8) | 0.375 |
| Charlson comorbidity index [median (IQR)] | 5 (3 - 6) | 4 (3 - 5) | **<0.0001** |
| Previous systemic corticosteroid therapy [n (%)] | 23 (12.7) | 5 (6.2) | 0.120 |
| Current or previous smoking [n (%)] | 69 (38.1) | 28 (35.0) | 0.630 |
| Symptoms at presentation [n (%)] |  |  |  |
| Cough | 144 (79.6) | 54 (67.5) | **0.036** |
| Fever | 140 (77.3) | 47 (58.8) | **0.002** |
| Dyspnoea | 122 (67.4) | 58 (72.5) | 0.412 |
| Diarrhoea | 55 (30.4) | 19 (23.8) | 0.273 |
| Myalgia | 53 (29.3) | 20 (25.0) | 0.477 |
| Expectoration | 48 (26.5) | 13 (16.2) | 0.071 |
| Impaired consciousness | 15 (8.3) | 4 (5.0) | 0.346 |
| NEWS at admission [median (IQR)]<sup>c</sup> | 3 (1 - 5) | 6 (3.5 - 7) | **<0.0001** |
| Interval from symptom onset to day 0, days<br>[mean ± SD]<sup>d</sup> | 10 (7 - 13) | 11 (9 - 16) | **0.001** |
| Interval from hospital admission to day 0, days<br>[mean ± SD]<sup>d</sup> | 4 (1 - 6) | 3 (1 - 8) | 0.432 |

CS: corticosteroids; IQR: interquartile range; NEWS: National Early Warning Score; SD: standard deviation; TCZ: tocilizumab.
<sup>a</sup> Includes chronic obstructive pulmonary disease, asthma, obstructive sleep apnea.
<sup>b</sup> Includes congestive heart failure and coronary heart disease.
<sup>c</sup> Data available for 236 patients.
<sup>d</sup> Day 0 refers to the date of administration of the first dose of immunomodulatory therapy (either corticosteroids or tocilizumab).

**Table 2.**
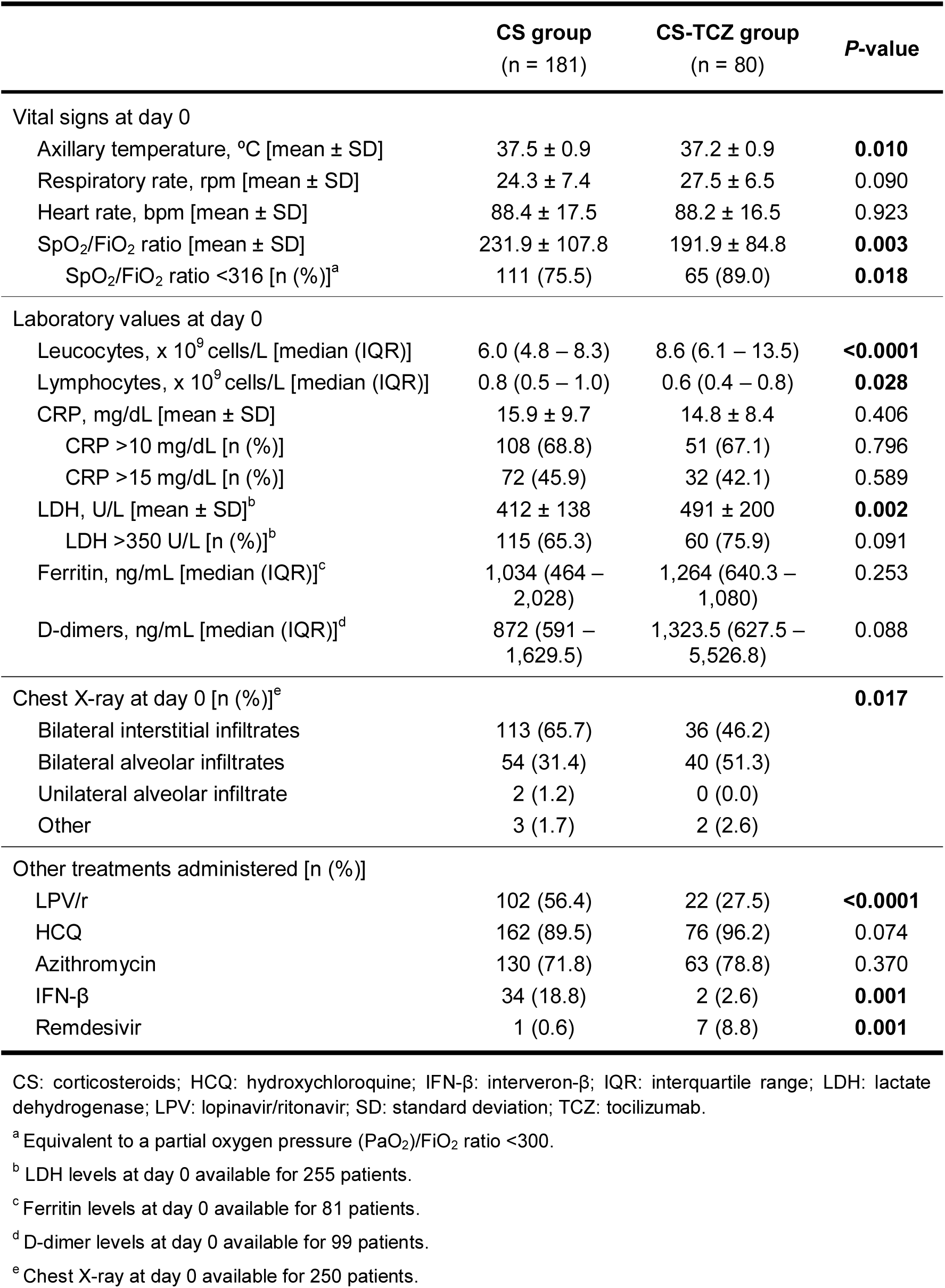
Vital signs and laboratory values at the initiation of immunomodulation, and other therapies used in both study groups.

Twenty-nine patients (11.1%) were admitted to the ICU at some point during the index hospitalization (5.5% [10/181] in the CS group and 23.8% [19/80] in the CS-TCZ groups). Immunomodulation was initiated prior to or on the same day of ICU admission in 48.3% (14/29) of these patients, within a median interval of 1 day (IQR: 0 - 5). In the overall cohort there were no differences between patients admitted or not admitted to the ICU in 14-day (27.6% [8/29] vs. 38.4% [89/232]; *P*-value = 0.258) or 28-day mortality rates (34.5% [10/29] vs. 42.2% [98/232], respectively; *P*-value = 0.424).

### Study outcomes

All-cause mortality by day +14 (primary outcome) was significantly lower in the CS-TCZ as compared to the CS group (22.5% [18/80] vs. 43.6% [79/181]; log-rank *P*-value <0.001) (**Figure 1a**). Regarding secondary outcomes, all-cause mortality by day +28 was also lower for patients within the CS-TCZ group (27.5% [22/80] vs. 47.5% [86/181]; log-rank *P*-value <0.001) (**Figure 1b**). A landmark analysis on those patients surviving ≥4 days (n = 208) confirmed the benefit survival by day +28 among patients in the CS-TCZ group (**Figure 2**). There were no significant differences in the rate of clinical improvement by day +14 (45.0% [36/80] vs. 35.4% [64/181] in the CS-TCZ and CS groups; *P*-value = 0.140).

**Figure 1:**
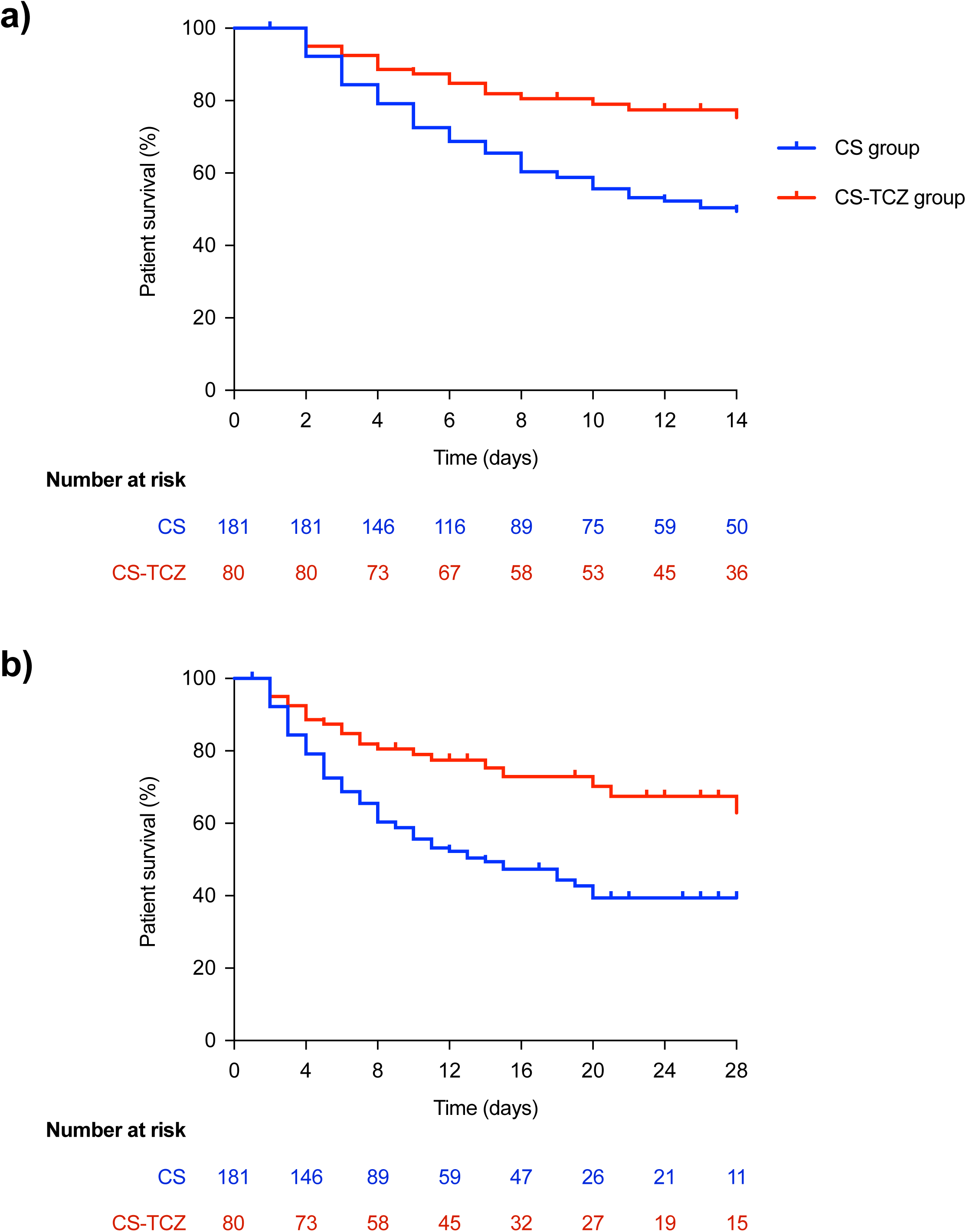
Comparison of Kaplan-Meier survival curves in patients included in the CS and CS-TCZ groups: **(a)** by day +14 (primary study outcome); **(b)** by day +28 (secondary study outcome). Log-rank *P*-values <0.001 for both comparisons. CS: corticosteroids; TCZ: tocilizumab.

**Figure 2.**
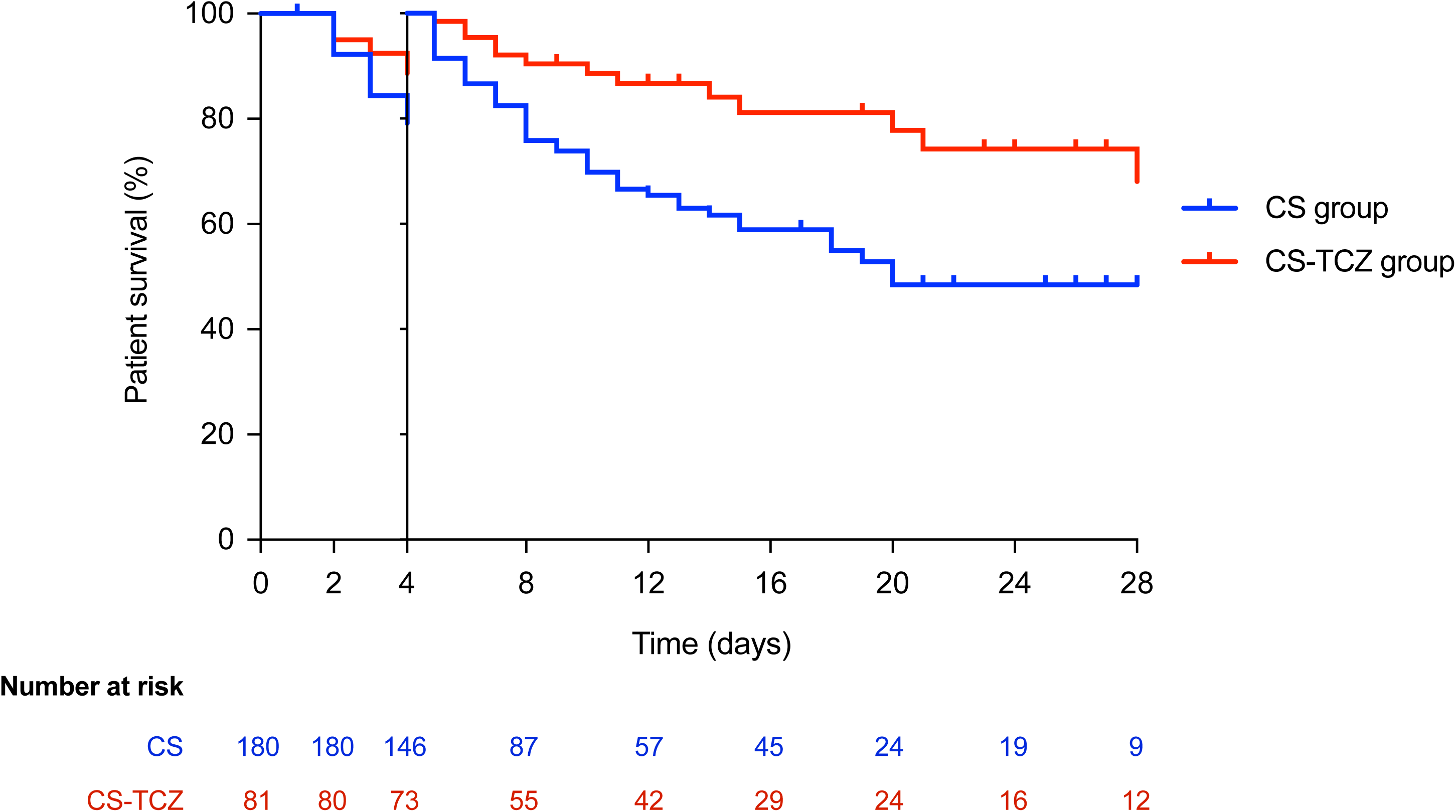
Landmark survival analysis on patients surviving by day +4. Log-rank test *P*-value = 0.0024. CS: corticosteroids; TCZ: tocilizumab.

### PS-adjusted analysis

In view of the presence of baseline imbalances between both groups, we constructed the PS for receiving TCZ associated to corticosteroids (CS-TCZ group) on the basis of those clinical and analytical variables present at day 0. The following measured predictors were included: age, Caucasian ethnicity, obesity, chronic lung disease, chronic heart disease, dementia, Charlson comorbidity index, cough at presentation, time since symptom onset, SpO_2_/FiO_2_ ratio <316 at day 0, lymphocyte count and LDH level at day 0, and the presence of bilateral alveolar infiltrates at day 0. The auROC of the PS was 0.849 (95% CI: 0.798 – 0.899), with a *P*-value = 0.128 in the Hosmer-Lemeshow test, suggesting excellent discriminative capacity and model calibration, respectively.

Patients in the CS-TCZ group experienced a lower 14-day all-cause mortality in the PS-adjusted model (HR: 0.34; 95% CI: 0.17 – 0.68; *P*-value = 0.002), after further adjustment for the receipt of antiviral agents (HR: 0.43; 95% CI: 0.25 – 0.74; *P*-value = 0.002), and in the IPTW-adjusted model (OR: 0.38; 95%: 0.21 – 0.68; *P*-value = 0.001). Regarding secondary outcomes, 28-day mortality was also significantly lower in the CS-TCZ group applying PS and IPTW adjustment, whereas clinical cure by day +7 was higher in the IPTW-adjusted model only (**Table 3**). These associations remained essentially unchanged in the landmark analysis restricted to hospitalized patients surviving at +4 (**Table S1** in Supporting Material).

**Table 3.**
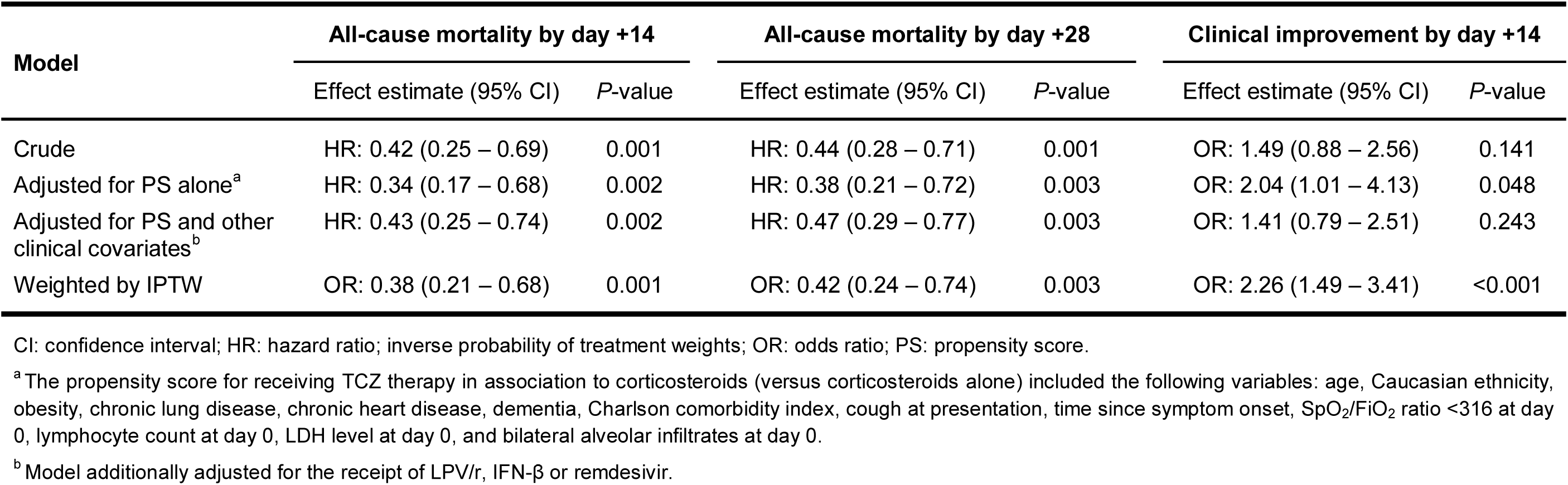
Effect of combination immunomodulatory therapy (corticosteroids plus TCZ) versus corticosteroids alone on primary and secondary study outcomes in different models.

| Model | All-cause mortality by day +14 |  | All-cause mortality by day +28 |  | Clinical improvement by day +14 |  |
| --- | --- | --- | --- | --- | --- | --- |
|  | Effect estimate (95% CI) | P-value | Effect estimate (95% CI) | P-value | Effect estimate (95% CI) | P-value |
| Crude | HR: 0.42 (0.25 – 0.69) | 0.001 | HR: 0.44 (0.28 – 0.71) | 0.001 | OR: 1.49 (0.88 – 2.56) | 0.141 |
| Adjusted for PS alone<sup>a</sup> | HR: 0.34 (0.17 – 0.68) | 0.002 | HR: 0.38 (0.21 – 0.72) | 0.003 | OR: 2.04 (1.01 – 4.13) | 0.048 |
| Adjusted for PS and other clinical covariates<sup>b</sup> | HR: 0.43 (0.25 – 0.74) | 0.002 | HR: 0.47 (0.29 – 0.77) | 0.003 | OR: 1.41 (0.79 – 2.51) | 0.243 |
| Weighted by IPTW | OR: 0.38 (0.21 – 0.68) | 0.001 | OR: 0.42 (0.24 – 0.74) | 0.003 | OR: 2.26 (1.49 – 3.41) | <0.001 |

CI: confidence interval; HR: hazard ratio; inverse probability of treatment weights; OR: odds ratio; PS: propensity score.
<sup>a</sup>The propensity score for receiving TCZ therapy in association to corticosteroids (versus corticosteroids alone) included the following variables: age, Caucasian ethnicity, obesity, chronic lung disease, chronic heart disease, dementia, Charlson comorbidity index, cough at presentation, time since symptom onset, SpO<sub>2</sub>/FiO<sub>2</sub> ratio <316 at day 0, lymphocyte count at day 0, LDH level at day 0, and bilateral alveolar infiltrates at day 0.
<sup>b</sup>Model additionally adjusted for the receipt of LPV/r, IFN-β or remdesivir.

### Treatment-related adverse events

The diagnosis of superinfection beyond day 0 was made in 6.1% (11/181) and 3.8% (3/80) of patients in the CS and CS-TCZ groups, respectively (*P*-value = 0.561). Hypertriglyceridemia (>200 mg/dL) was more common in the CS-TCZ group (18.8% [15/80] vs. 0.0% [0/181]; *P*-value <0.0001). Elevation in liver function tests was observed in 3.9% (7/181) and 8.8% (7/80) of patients in the CS and CS-TCZ groups, respectively (*P*-value = 0.136).

## Discussion

An increasing number of studies (including RCTs) supports the use of systemic corticosteroids to reduce mortality in severe forms of COVID-19 (7), although the incremental effect of adding TCZ to this therapeutic approach remains unclear. The shortage of TCZ at the initial stages of the pandemic in our setting offered the opportunity to compare two consecutive cohorts of patients ≥65 years treated with different immunomodulation strategies. Patients in the CS group were older and had a greater comorbidity burden than those in the CS-TCZ group, a difference that may reflect the reluctance of treating physicians to initiate IL-6 blockade in this aged population due to concerns about potential adverse events and the greater clinical experience with the use of short courses of corticosteroids. Indeed, only two patients older than 90 years underwent combination therapy, whereas 8.3% of those in the CS group were in this age stratum. Another difference was the proportion of patients admitted to the ICU during the index hospitalization (5.5% and 23.8% in the CS and CS-TCZ groups), which is likely due the evolving availability of ICU resources throughout the study period, with less beds available at the peak of the pandemic in March 2020 in coincidence with the shortage of TCZ. Nevertheless, we found not differences in mortality between patients admitted or not admitted to the ICU.

Combination therapy with corticosteroids and TCZ was found to be associated in the present cohort with lower mortality rates at 14 and 28 days as compared to the use of corticosteroids alone. The baseline imbalances in terms of age and comorbidities mentioned above would have favored the CS-TCZ group, although the clinical status at day 0 was worse in these patients (as reflected by their lower SpO_2_/FiO_2_ ratio and lymphocyte count, higher LDH level, more common bilateral alveolar involvement in the chest X-ray, and longer interval from symptom onset). The experimental use of remdesivir —a direct-acting antiviral agent inhibiting the SARS-CoV-2 RNA-dependent RNA polymerase that has been demonstrated to shorten the time to recovery (24)— was more common in the CS-TCZ group (albeit it was administered to only 8.2% of patients). In addition, more patients in the CS group received IFN-β, in keeping with the recommendations contained in the institutional protocol during the first weeks of March. Therefore, we applied PS adjustment and IPTW to take into account these differences operating in opposite senses, and still found significant risk reductions in the primary study outcome ranging from 57% to 66% (according to the adjusted model) in favor of the combination of corticosteroids and TCZ. On the other hand, and despite the lack of apparent impact in the crude comparison, the odds for achieving clinical response by day +14 was also significantly higher in the CS-TCZ group after balancing potential confounders by IPTW. Nevertheless, this association should be taken with caution since it was not confirmed in PS-adjusted models.

Although it was to be expected that the old age of our patients (77.2 ± 7.7 years) would have put them at an increased risk of TCZ-associated adverse events, no significant differences were found in the occurrence of bacterial or fungal infectious complications between study groups. No apparent impact on the risk of secondary bacterial infection among patients treated with TCZ and corticosteroids (versus no immunomodulatory therapy) was reported from a recently published multicenter study either(25). The increase of lipid levels is a well-known event in patients with rheumatoid arthritis receiving long-term TCZ therapy(26), and the development of TCZ-induced hypertriglyceridemia has been also observed in the setting of COVID-19(27).

Only few previous studies have investigated the effectiveness of the association between TCZ and corticosteroids for severe COVID-19 pneumonia. Hazbun et al. described a small series of patients undergoing mechanical ventilation with no control group (28). Ramiro et al. compared a two-step approach based on high-dose methylprednisolone followed by TCZ if needed in non-responders with a historic control group receiving standard of care alone. The authors found that this sequential strategy accelerated respiratory recovery, decreased in-hospital mortality and reduced the likelihood of invasive mechanical ventilation (29). On the contrary, Rodríguez-Baño et al. reported no differences in the risk of intubation or death among patients treated with combination therapy(25).

There are a number of limitations in our study to be acknowledged. First, due to its retrospective observational design, the impact of unmeasured confounders cannot be completely ruled out, even after PS-based and IPTW adjustments. As discussed above, the direction of potential bias is not obvious in view of the nature of baseline imbalances. Likewise, we have attempted to minimize the risk of survivor bias by excluding those patients dying within the first 24 hours and by means of landmark survival analyses. Second, different regimens of corticosteroids and TCZ were used, preventing us to conclude on optimal dosing strategies. Finally, the single-center design hampers the applicability of findings to other settings.

The Infectious Disease Society of America has recently highlighted in their evidence-based guidelines (19) that RCTs constitute the optimal framework to evaluate the efficacy and safety of newer therapies for COVID-19. Indeed, a number of ongoing trials are aimed at comparing TCZ versus corticosteroids (NCT04345445 and NCT04377503), corticosteroids alone or associated to TCZ (NCT04476979), and TCZ versus TCZ plus corticosteroids (NCT04486521). Until the results of these RCTs become available, observational studies might be useful to guide clinical decisions providing that caution is applied before concluding on potential causation. In conclusion, the present experience suggests that the use of IV TCZ on top of corticosteroid therapy may be more useful than corticosteroids alone to improve outcomes in patients ≥65 years with hyperinflammatory status triggered by SARS-CoV-2 infection.

## Data Availability

Available under request

## Appendix

### Other members of the H12O Immunomodulation Therapy for COVID-19 Group

Unit of Infectious Diseases: Manuel Lizasoain, Tamara Ruiz-Merlo, Patricia Parra; Department of Pharmacy: José Miguel Ferrari; Department of Pneumology: Javier Sayas Catalán, Eva Arias Arias; Department of Nephrology: Fernando Caravaca, Amado Andrés, Manuel Praga; Department of Rheumatology: María Martín-López; Department of Hematology: Denis Zafra, Cristina García Sánchez; Department of Oncology: Carmen Díaz-Pedroche, Flora López, Luis Paz-Ares; Department of Intensive Care Medicine: Jesús Abelardo Barea Mendoza, Paula Burgueño Laguía, Helena Domínguez Aguado, Amanda Lesmes González de Aledo, Juan Carlos Montejo; Department of Emergency Medicine: Antonio Blanco Portillo, Laura Castro Reyes, Manuel Gil-Mosquera, José Luis Montesinos Díaz, Isabel Fernández-Marín; Department of Immunology: Óscar Cabrera-Marante, Antonio Serrano-Hernández, Daniel Pleguezuelo, Édgar Rodríguez de Frías, Paloma Talayero, Laura Naranjo-Rondán, Ángel Ramírez-Fernández, María Lasa-Lázaro, Daniel Arroyo-Sánchez, Rocío Laguna-Goya; Department of Microbiology: Rafael Delgado, María Dolores Folgueira.

### Transparency declaration

All the authors declare no potential conflict of interest regarding this study.

## Acknowledgements

The authors would like to acknowledge all the health-care workers involved in the response to the current pandemic in our hospital and, singularly, those who suffered COVID-19.

## Supporting Material

### Supplementary Results

- Table S1. Effect of combination immunomodulatory therapy (corticosteroids plus TCZ) versus corticosteroids alone on primary and secondary study outcomes in the landmark analysis of surviving patients that remained hospitalized by day +4 (n = 208).

